# Rapid increase of SARS-CoV-2 variant B.1.1.7 detected in sewage samples from England between October 2020 and January 2021

**DOI:** 10.1101/2021.03.03.21252867

**Authors:** Thomas Wilton, Erika Bujaki, Dimitra Klapsa, Martin Fritzsche, Ryan Mate, Javier Martin

## Abstract

SARS-CoV-2 variants with multiple amino acid mutations in the spike protein are emerging in different parts of the world raising concerns on their possible impact on human immune response to the virus and vaccine efficacy against them. Recently, a variant named lineage B.1.1.7 was detected and shown to be rapidly spreading across the UK since November 2020. As surveillance for these SARS-CoV-2 variants of concern (VOCs) becomes critical, we have investigated the use of environmental surveillance (ES) for the rapid detection and quantification of B.1.1.7 viruses in sewage as a way of monitoring its expansion that is independent on the investigation of identified clinical cases. B.1.1.7 mutations in viral sequences from sewage were first identified in a sample collected in London on 10^th^ November 2020 and shown to rapidly increase in frequency to >95% in January 2021, in agreement with clinical data over the same period. We show that ES can provide an early warning of VOCs becoming prevalent in the population and that, as well as B.1.1.7, our method can potentially detect VOCs B.1.351 and P.1, first identified in South Africa and Brazil, respectively, and other viruses also carrying critical spike mutation E484K, known to have an effect on virus antigenicity.

## Introduction

A novel SARS-CoV-2 lineage B.1.1.7 was recently identified and shown to be rapidly expanding across the UK, particularly in London, the South East, and the East of England [1, 2]. This lineage was detected in November 2020, and likely originated in September 2020 in Kent, South East England. B.1.1.7 viruses possess a striking constellation of nucleotide changes with an unusual number of non-synonymous substitutions in the spike protein of potential biological relevance [3] (Fig. 1). For this reason, lineage B.1.1.7, was designated SARS-CoV-2 Variant of Concern (VOC) 202012/01 by Public Health England (PHE) as the mutations found might impact transmission, immune response and disease severity. Mutation N501Y, located within the receptor-binding domain (RBD), can increase binding affinity to human and murine angiotensin-converting enzyme (ACE)-2 receptor and cell infectivity in mice [4-6]. Substitution P681H is immediately adjacent to the furin cleavage site, essential for viral infectivity [7, 8], while deletion of amino acids 69-70 has been identified in multiple lineages associated with different RBD mutations and has been related to the evasion to human immune response [9]. The deletion of nucleotides 21765-21770 coding for amino acids 69-70 has been associated with S-gene target failure (SGTF) in diagnostic PCR testing of samples showing positive results for other gene targets [1] and is used to estimate the proportion of COVID-19 confirmed cases likely associated with infection by variant B.1.1.7. Epidemiological evidence and phylogenetic analysis of viral sequences suggest that viruses of the B.1.1.7 lineage may have increased transmissibility compared with ancestral isolates [2]. Whole-genome sequencing and SGTF data have shown a rapid increase in the prevalence of B.1.1.7 variant through time with the proportion of sequences from clinical samples associated with B.1.1.7 in England reaching around 94% during the last week of January 2021 [1, 2, 10]. Additional analyses indicate exponential growth of lineage B.1.1.7 and an increase of between 0.4 and 0.7 in the reproduction number with respect to previous lineages [2].

**Figure 1.**
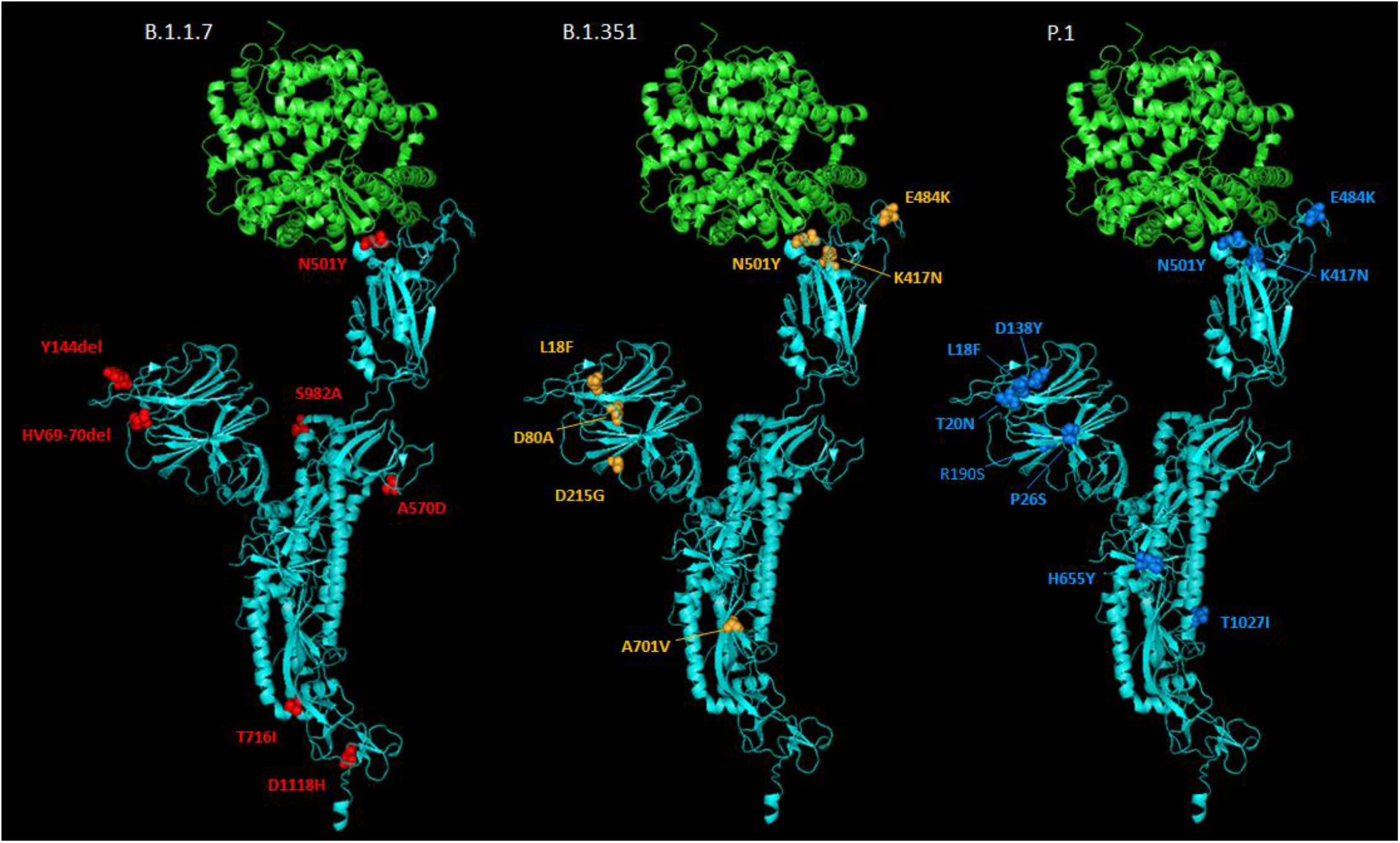
Molecular cartoon diagram of the three-dimensional structure of SARS-CoV-2 spike protein monomer in the open form (cyan) in complex with receptor ACE-2 (green). The image was generated using PyMOL Molecular Graphics System, Version 1.7.0.3 software (Schrödinger, LLC) using cryo-EM data (Protein Data Bank accession number 7DF4.pdb [31]). Amino acid substitutions found in B.1.1.7 (red), B.1.351 (orange) and P.1 (blue) variants are shown.

Other VOCs have been identified recently, notably lineages B.1.351 and P.1, known to be circulating in South Africa since early October 2020 [11] and in Brazil since early December 2020 [12], respectively, and also containing complex mutation constellations including N501Y amino acid change (Fig. 1). B.1.351 and P.1 variants have two more spike mutations of biological significance: E484K and K417N/T, both located in the RBD and both important for ACE-2 binding and antibody recognition. Mutations at spike amino acid 484 have been shown to reduce binding to convalescent serum antibodies with neutralization by some sera reduced >10-fold [13]. Amino acid substitutions at residue 417, K417N in B.1.351 and K417T in P.1 appear to improve evasion from antibodies in combination with N501Y and E484K [14]. More recently, mutation E484K has been identified in a small number of B.1.1.7 viruses in England [15], raising increased concerns about this variant. The effects of mutations found in the VOCs detailed above suggest that VOCs could be a threat to the protective efficacy of current vaccines by reducing vaccine-induced immunity against them. Recent research has shown that while B.1.1.7 is modestly resistant to sera from vaccinees, B.1.351 is markedly more resistant to neutralization by such sera and some COVID-19 vaccines appear to show reduced efficacy against this variant [16-20]. Although further research is required to fully understand the effect of spike mutations on vaccine efficacy, updating vaccine strains might be required in the future in a similar manner to what is common practice for flu vaccines.

The appearance and rapid growth of VOCs shows the need for enhanced genomic and epidemiological surveillance worldwide to ensure that any new spike changes that might arise are rapidly detected and their biological effects investigated. Environmental surveillance (ES) has proven to be a sensitive method for the detection and monitoring of SARS-CoV-2 circulation as described in multiple reports from many countries (reviewed in [21]). As ES can produce a real-time snapshot of virus transmission at a given time point, including that from asymptomatic infections, it might provide precise information on the prevalence of VOCs in the population. We were able to detect SARS-CoV-2 viral RNA in sewage samples collected in London during the first wave of the pandemic with variation in viral RNA levels comparing to that of numbers of confirmed COVID-19 cases [22]. In addition, using next generation sequencing (NGS) analysis of DNA amplicons generated from sewage concentrates, we were able to track changes in variant predominance during the first stages of the pandemic detecting variants that were particularly prevalent in the UK and the rapid expansion of D614G variant reaching nearly complete dominance in May 2020 [22]. This variant, named D614G because it contains a mutation from A to G at nucleotide 23,403 resulting in amino acid change from Aspartic acid to Glycine at residue 614 of the spike protein, appeared to increase viral infectivity and transmissibility and was firstly described in late February 2020 in Italy rapidly expanding and becoming the dominant SARS-CoV-2 variant globally few months later [23]. Similar work by other groups have described the detection of co-circulating SARS-CoV-2 variants in sewage samples from different locations including a recent report describing the identification of B.1.1.7 lineage in sewage samples from Switzerland [24-28].

In order to further evaluate the value of ES for SARS-CoV-2 detection, we have analysed sewage samples collected in London between 14^th^ January 2020 and 26^th^ January 2021 for the presence of SARS-CoV-2 using a semi-quantitative nested RT-PCR (nPCR) system targeting two different genomic regions as we have found this system to be more sensitive and consistent than quantitative RT-PCR assays [22]. More importantly, we have designed new nPCR reactions to generate DNA amplicons and analyse them by NGS with an aim to specifically detect and quantify the presence of key mutations that discriminate lineages B.1.1.7, B1.351 and P.1 from ancestral isolates and between themselves.

## Methods

### Wastewater sample collection and processing

One-litre inlet wastewater composite samples were collected during a 24-hour period at a sewage plant in London with a catchment area of approximately 4.0 x 10^6^ people. Monthly samples collected every second Tuesday of the month were collected between 14^th^ January 2020 with an additional sample collected on 26th January 2021. Each sample was processed using a filtration-centrifugation method described before [22]. Briefly, following removal of solids by centrifugation at 3,000g, raw sewage was filtered through a 0.45 µM filter (Nalgene 500 ml Rapid-flow™) and concentrated down to 200-400 μl using Merck Millipore Centricon™ Plus-70 Centrifugal Filter Units with a 10 kDa molecular weight cut-off (Merck), following manufacturer’s instructions. Samples from January to May 2020 reported before [22] were re-analysed in this study.

### nPCR amplifications

Viral RNA was purified from sewage concentrates using the High Pure viral RNA kit (Roche). RT-PCR fragments were amplified from purified viral RNAs using an nPCR system described before consisting of a one-step RT-PCR reaction followed by a second PCR using the first PCR product as template [22]. Genome location and nucleotide sequences of primer sets used for the PCR reactions are shown in Table S1. Primer sets A and B targeting RdRP and ORF8b gene regions were used for diagnostic nPCRs. Primer sets C and D targeting spike gene regions were used for the detection of B.1.1.7, B.1.351 and P.1 lineages. As shown in Table 1, sequence analysis of nPCR C and D products can discriminate between B.1.1.7, B.1.351 and P.1 lineages and between them and ancestral isolates. Primers were tested using serial dilutions of purified RNA from NIBSC’s virus reagent 19/304 containing non-infectious synthetic SARS-CoV-2 RNA packaged within a lentiviral vector (https://www.nibsc.org/products/brm_product_catalogue/detail_page.aspx?catid=19/304). 12 RNA aliquots from each sewage sample were used for nPCR amplifications for each nPCR reaction. Good laboratory practices were observed in all assays to reduce the possibility of cross-contamination; using different laboratory locations for sample processing, preparation of reaction mixtures, template addition and post-processing analysis. RNA extraction and negative template controls were included in every assay. Selected purified DNA amplicon products were sequenced by the Sanger method using an ABI Prism 3130 genetic analyser (Applied Biosystems).

**Table 1.**
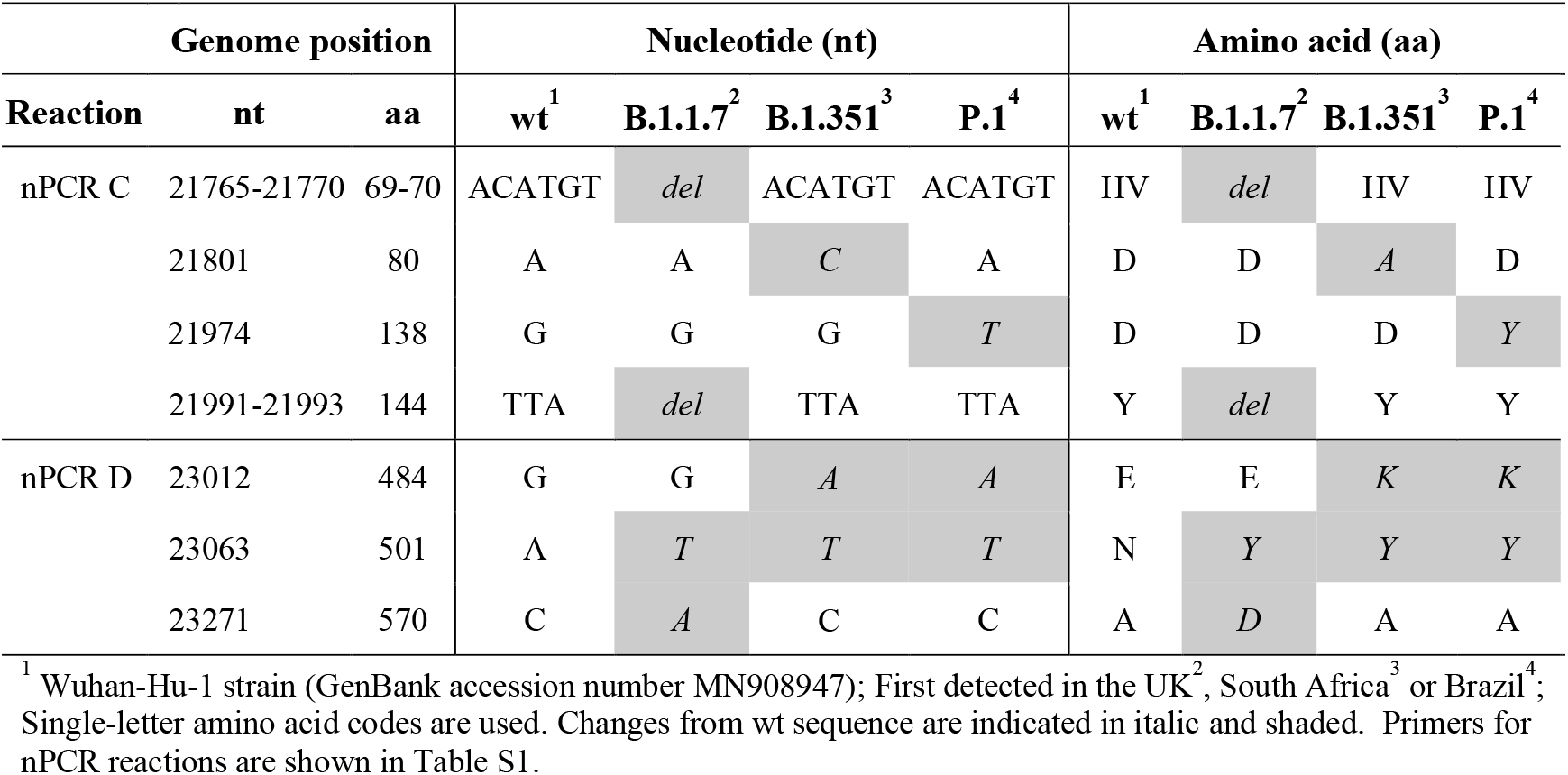
Nucleotide and amino acid differences between VOCs and ancestral isolates

### NGS analysis of nPCR products

NGS libraries were constructed as reported before [22] using the DNA Prep kit (formerly known as Nextera DNA Flex) and dual-indexed using Nextera DNA CD Indexes (both Illumina). These libraries were pooled in equimolar concentrations and sequenced with 250 bp paired-end reads on MiSeq v2 (500 cycles) kits (Illumina, USA). Initial demultiplexing was performed on-board by the MiSeq Reporter software. FASTQ sequencing data was adapter and quality trimmed by Cutadapt v2.10 [29] for a minimum Phred score of Q30, minimal read length of 75 bp, and 0 ambiguous nucleotides.

### Generation of SARS-CoV-2 sequence contigs and identification of SNPs

Further processing and analysis of NGS data was performed using Geneious 10.2.3 software as described before [22]. Filtered reads were imported into Geneious 10.2.3, paired end reads combined and merged and sequence contigs built by reference-guided assembly. Reads were mapped to references with a minimum 50 base overlap, minimum overlap identity of 90%, maximum 10% mismatches per read, allowing up to 15% gaps, and index word length of 12. Single nucleotide polymorphisms (SNPs) were identified using Geneious 10.2.3 default settings. The original SARS-CoV-2 Wuhan-Hu-1 strain (GenBank accession number MN908947) was used as a reference. Variants with coverage <500, average quality <30, variant frequency <5% and variant P-value >10^−6^ were excluded. The analysis of several amplicon replicates per sample is critical to reduce sampling effects and increase the accuracy of SNP determinations for viral genomes present in a given sewage sample. If we set up a conservative limit of 5% for any SNP found in any replicate amplicon sequence to be accepted as genuinely present in viral RNAs, and given that we sequenced 12 independent RNA replicates per sample and nPCR reaction, the limit of detection for detecting a sequence variant would be approximately 0.42%, which seems reasonably low for the early detection of the spread of SARS-CoV-2 variants of potential concern. If we consider sequencing 5% of all positive clinical samples as a minimum requirement, which is currently only achieved in a handful of countries, a 0.42% prevalence of a variant would mean finding 21 to 105 viruses among 10,000 to 50,000 new daily positive cases, respectively. However, for clinical surveillance to be representative and have comparable sensitivity to ES, widespread and consistent clinical sampling across the sewage catchment area would be needed.

### SARS-CoV-2 nucleotide sequence analysis and estimates of the prevalence of B.1.1.7 lineage among whole-genome sequences from clinical samples

SARS-CoV-2 sequences obtained in this study were compared to those available in the Global Initiative on Sharing All Influenza Data (GISAID) SARS-CoV-2 sequence database [10]. The COVID-19 Genomics (COG-UK) Consortium / Mutation Explorer with sequencing data generated by 28th February 2021 (http://sars2.cvr.gla.ac.uk/cog-uk/) was also used to examine the presence and proportion of selected viral amino acid variations in clinical samples. Geneious version 10.2.3 software (Biomatters) was used for these analyses. Estimates of the proportion of viruses belonging to B.1.1.7 lineage at different time points in England were obtained using the GISAID SARS-CoV-2 sequence database [10] with results available on 1^st^ March 2021.

### Statistical analysis

Statistical analysis and graphical representation of sequence frequency data were conducted using GraphPad Prism version 8.1.2 software.

## Results

### nPCR amplifications to estimate SARS-CoV-2 RNA levels in sewage samples

Twelve different RNA replicates from each sewage concentrate were used to generate nPCR products with primer combinations A and B (Table S1) targeting RdRP and ORF8b gene regions, respectively. 14 sewage samples collected between 14^th^ January 2020 and 26^th^ January 2021 were analysed. The proportion of positive PCR results for each sample was used as a semi-quantitative comparison of RNA levels present in sewage samples. The results are shown in Fig. 2 in the context of epidemiological data. As we have shown before, a sample from 14^th^ January 2020 was negative and only low levels of viral RNA were detected in the sample from 11^th^ February 2020 [22], 3 days before the first COVID-19 cases were confirmed in the area. Viral RNA concentration increased in March and April during the first wave of the pandemic and sharply declined in May as a result of nationwide lockdown measures introduced from 23rd March. Following a period of about 4 months with low level virus transmission, SARS-CoV-2 RNA levels increased again in sewage from early September in agreement with increasing COVID-19 confirmed cases. Selected PCR products from all sewage concentrates were analysed by Sanger sequencing confirming their SARS-CoV-2 sequence.

**Figure 2.**
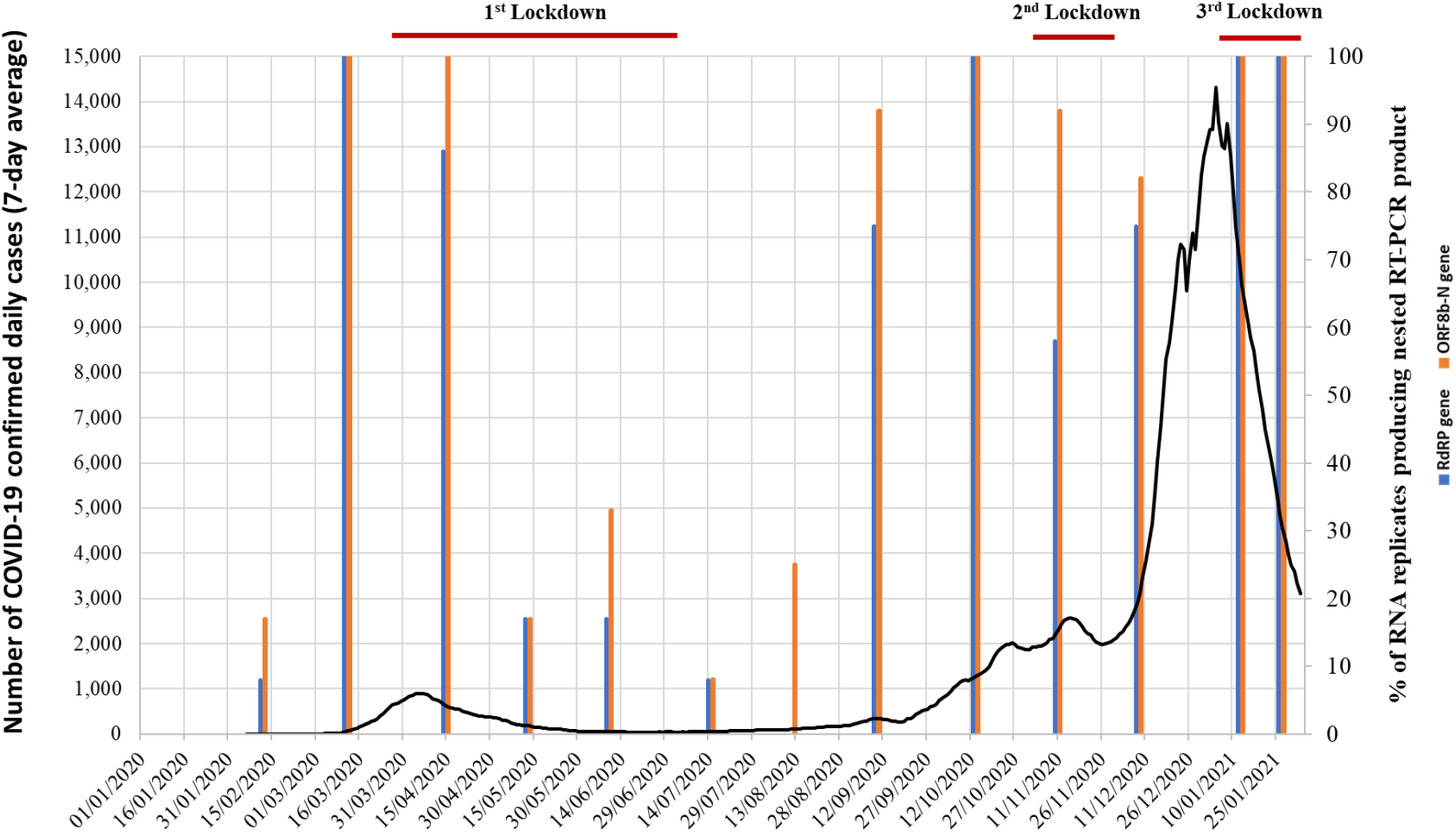
Detection of SARS-CoV-2 RNA in sewage samples collected in London between 14^th^ January 2020 and 26^th^ January 2021. The % of RNA replicates producing positive results in nPCR A (blue) and nPCR B (orange) reactions targeting RdRP and ORF8b genomic regions, respectively, are shown as vertical columns. The number of daily confirmed COVID-19 cases in London are represented with a black line (7-day rolling average). Nationwide lockdown periods are indicated as horizontal red lines. Source for COVID-19 cases data: https://coronavirus.data.gov.uk/

### NGS analysis of nPCR products for the detection of SARS-CoV-2 B.1.1.7, B1.351 and P.1 lineages

Twelve additional RNA replicates from each sewage concentrate were used to generate nPCR products with primer combinations C and D (Table S1), mapping in the spike protein gene. All amplicons from positive nPCR reactions were analysed by NGS with an aim to detect and quantify nucleotide sequence variations specifically found in B.1.1.7, B1.351 and P.1 lineages (Table 1). Nucleotide sequences characteristic of lineage B.1.1.7 were detected in sewage samples from 10^th^ November 2020 onwards. Mean sequence frequency values at key nucleotide positions, calculated from all sequenced replicates for each monthly sewage sample, are shown in Fig. 3. The results show that mutations discriminating B.1.1.7 lineage from ancestral isolates, which include relevant mutations coding for amino acid changes and amino acid deletions (Table 1), clearly increased in frequency between November 2020 and January 2021 from 6.84-8.88% on 10^th^ November to 43,99-46.36% on 8^th^ December 2020, 93.60-94.13% on 12^th^ January 2021 and 95.25-98.83% on 26^th^ January 2021. An example of NGS results showing an increasing proportion of nucleotides 21765–21770 deletion (deletion of amino acids HV69-70) in viral RNAs from sequential sewage samples is shown in Fig. S1. Estimates of B.1.1.7 prevalence based on NGS data were in very good agreement with those from whole-genome sequencing and SGTF data from clinical samples from London and England [1, 2]; according to the GISAID sequence database [10], the frequency rates of B.1.1.7 sequences from England during the week before and the week after sewage sample collection were 3.42% and 7.34% for 10^th^ November, 25.19% and 42.67% for 8^th^ December, 81.08% and 83.44% for 12^th^ January, and 89.61% and 94.26% for 26^th^ January, very much in line to what was observed in sewage (Table 2). As shown in Fig. 4, frequency rates of B.1.1.7 mutations found in viral RNAs from sewage and those of B.1.1.7 whole-genome sequences among clinical samples in England superimpose almost perfectly when represented together (using clinical data from one week after sewage sample collection dates). In contrast, none of the nucleotide substitutions unique to B.1.351 and P.1 lineages (Table 1) in any of the three nPCR products were detected in relevant amplicons from any of the sewage samples.

**Table 2.** Frequency of SARS-CoV-2 B.1.1.7 lineage in sewage and clinical samples

|  |  |  | **B.1.1.7 mutations (%)** |  |  |  |
| --- | --- | --- | --- | --- | --- | --- |
| **Sample** |  | **Date** | **21765-21770**<br>**deletion** | **21991-21993**<br>**deletion** | **A23063T** | **C23271A** |
| Oct | ES | 13.10.20 | 0.00 | 0.00 | 0.00 | 0.00 |
|  | Clinical | 06-12.10.20 | 0.08 | 0.08 | 0.08 | 0.08 |
|  | Clinical | 13-19.10.20 | 0.06 | 0.06 | 0.06 | 0.06 |
| Nov | ES | 10.11.20 | 8.53 | 8.88 | 6.88 | 6.84 |
|  | Clinical<sup>1</sup> | 03-09.11.20 | 3.42 | 3.42 | 3.42 | 3.42 |
|  | Clinical | 10-16.11.20 | 7.34 | 7.34 | 7.34 | 7.34 |
| Dec | ES | 8.12.20 | 43.99 | 44.34 | 46.36 | 46.28 |
|  | Clinical<sup>1</sup> | 01-07.12.20 | 25.19 | 25.19 | 25.19 | 25.19 |
|  | Clinical | 08-14.12.20 | 42.67 | 42.67 | 42.67 | 42.67 |
| Jan.1 | ES | 12.01.21 | 94.13 | 93.9 | 93.86 | 93.6 |
|  | Clinical | 05-11.01.21 | 81.08 | 81.08 | 81.08 | 81.08 |
|  | Clinical | 12-18.01.21 | 83.44 | 83.44 | 83.44 | 83.44 |
| Jan.2 | ES | 26.01.21 | 98.83 | 98.78 | 95.3 | 95.25 |
|  | Clinical | 19-25.01.21 | 89.61 | 89.61 | 89.61 | 89.61 |
|  | Clinical | 26.01.21 to<br>01.02-21 | 94.26 | 94.26 | 94.26 | 94.26 |

Cells are shaded according to values shown, from the lowest (light grey) to the highest (dark grey).
<sup>1</sup> Data from the GISAID sequence database [10] consulted on 13 February 2021.

**Figure 3.**
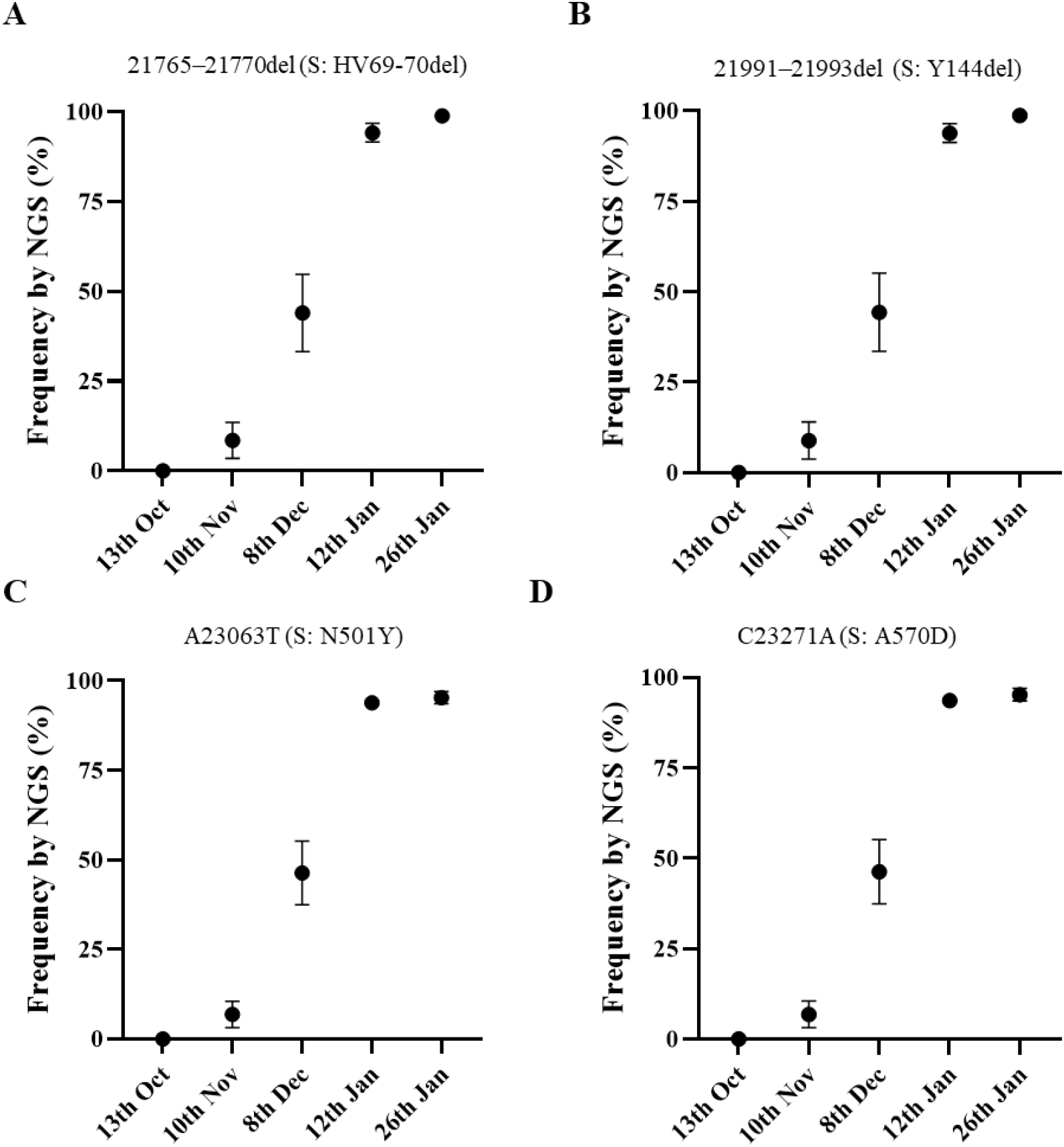
Mean frequency values of nucleotide mutations in the spike gene distinctive of SARS-CoV-2 B.1.1.7 lineage identified in nPCR C and nPCR D products synthesized from sewage concentrates and measured by NGS analysis. A) deletion of nucleotides 21765–21770 (deletion of amino acids HV69-70); B) deletion of nucleotides 21991–21993 (deletion of amino acid Y144); C) mutation A23063T (amino acid change N501Y); and D) mutation C23271A (amino acid change A570D). Error bars indicate standard error of the mean.

**Figure 4.**
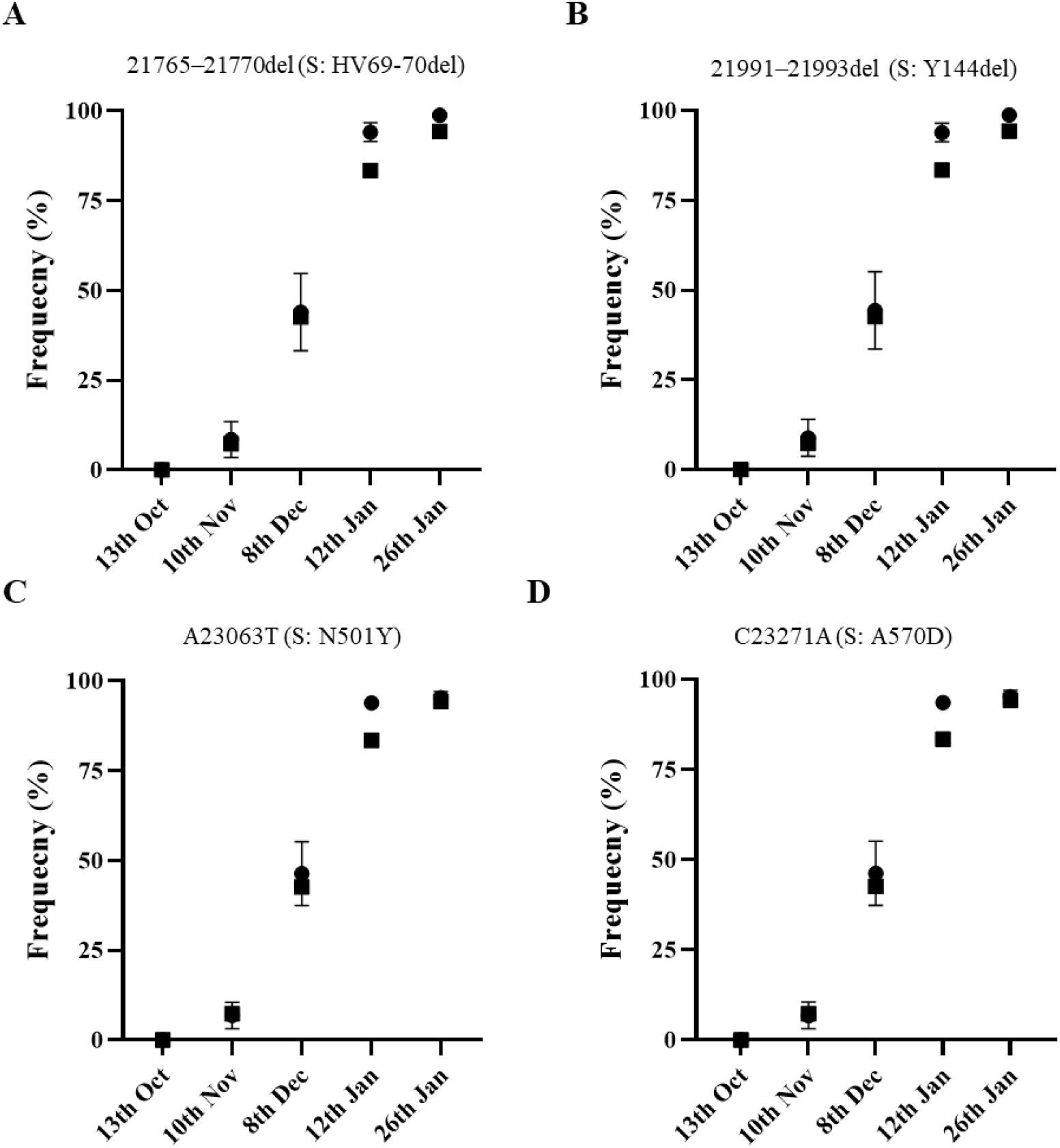
Frequency rates of B.1.1.7 whole-genome sequences among clinical samples from England collected during the week immediately after sewage sample collection are shown as black squares superimposed to mean frequency values of B.1.1.7 mutations found in sewage samples measured by NGS (black circles with error bars indicating standard error of the mean, as shown in Fig. 3).

Additional nucleotide sequence variations were found at different sites in amplicons from sewage samples, but none followed the same pattern as that observed with B.1.1.7 mutations, which were repeatedly detected in replicate viral RNA sequences and consistently found in sequential samples in increasing proportions.

## Discussion

We were able to detect SARS-CoV-2 RNA in sewage samples from London throughout the COVID-19 pandemic with changes in viral RNA levels shown to be in good agreement with those of confirmed COVID-19 cases reflecting national restrictions (Fig. 2). The second lockdown period lasting from 5^th^ November to 2^nd^ December appear to only have had a limited effect in reducing virus transmission which, together with the emergence of the apparently highly transmissible B.1.1.7 variant, may explain the high levels of viral RNA found in sewage and the high number of SARS-CoV-2 infections reported in England in December 2020 and January 2021. One in 50 people in England (1 in 30 in London) were estimated to have had SARS-CoV-2 infection in the week ending 2 January according to an infection survey by the Office for National Statistics [30]. The third nationwide lockdown initiated on 4th January 2021 is having a clear effect at reducing virus transmission as judged by the observed drop in COVID-19 confirmed cases, but as of 26^th^ January 2021, SARS-CoV-2 RNA levels remained high in sewage (Fig. 2) reflecting the extent of virus transmission occurring during the third wave of the COVID-19 epidemic. Although, detailed quantification of SARS-CoV-2 RNA in sewage and its correlation with COVID-19 infection prevalence was outside the scope of this paper and would require more frequent and widespread sampling, our results suggest that high viral RNA concentration levels could be detected preceding peaks in COVID-19 confirmed cases showing the potential predictive value of ES as it has been shown in other studies [21].

NGS analysis of DNA amplicons directly synthesized from SARS-CoV-2 RNA present in sewage concentrates from London allowed us to readily identify viruses belonging to B.1.1.7 lineage before widespread transmission of this variant was obvious. We were able to detect B.1.1.7 variant at a frequency of 6.84-8.88% in a sample collected on 10^th^ November 2020, few weeks before it was first noticed through clinical surveillance. B.1.1.7 variant first became apparent in late November 2020 when PHE was investigating why infection rates in Kent remained high despite national restrictions. A concerning cluster of virus isolates showing an unusual constellation of mutations was then found that could explain the persistent high infection rates [3, 15]. Retrospective analysis showed that the two earliest genomes belonging to B.1.1.7 lineage were collected on 20^th^ September and 21^st^ September 2020 in Kent and London, respectively. B.1.1.7 is believed to have circulated at very low levels in the population until mid-November from when it rapidly spread across the country. The frequency rates of B.1.1.7 sequences observed in sewage between November 2020 and January 2021 were very similar to those of COVID-19 cases associated with infection by B.1.1.7 viruses at around the time of sewage sample collection (Table 2). B.1.1.7 was not detected in the sewage sample from 13^th^ October 2020 consistent with the fact that the frequency of B.1.1.7 viruses identified in clinical samples collected in the weeks before and after 13^th^ October 2020 was <0.08% in the whole of England. The proportion of B.1.1.7 sequences found in sewage increased rapidly from early November reaching >90% eight weeks after the first detection in agreement with increasing infections associated with B.1.1.7. No evidence of the presence of variants B.1.351 and P.1 was found in any of the sewage samples, which was expected as only a small number of B.1.351 isolates had been found in England at that time, not necessarily in London, and no P.1 viruses had been detected in the UK yet. G23012A nucleotide substitution present in both B.1.351 and P.1 lineages and responsible for spike amino acid change E484K and which is thought to play a critical role at changing virus antigenicity and reducing human immune response to the virus and vaccine efficacy [13, 16-20], was not detected in viral genomes from sewage. This mutation has recently been found in a small number of B.1.1.7 isolates in England but mostly in locations away from London [15]. We expect that the methods shown here can help to rapidly identify such mutants as well as B.1.351 and P.1 lineages in the same way that we have identified B.1.1.7 viruses (Table 1). These methods can also be used to sequence SARS-CoV-2 in clinical samples using traditional Sanger sequencing for the rapid identification of VOCs in settings were whole-genome sequencing capabilities are limited. New PCRs can be quickly designed to track down new VOCs that might arise anywhere and detect importations. Furthermore, systematic sequencing of the spike gene of viral RNAs present in sewage samples collected regularly would help proactively identifying potential new VOCs based on mutations and trends encountered. Eventually, if vaccine strains need to be regularly updated as it is current practice with flu vaccines, ES could be a useful resource to help vaccine strain selection based on prevalent SARS-CoV-2 variants circulating around the world.

As we show here, ES has a potential advantage for the early detection of VOCs versus clinical surveillance as it provides an immediate snapshot of virus transmission events representing thousands/millions of people, including asymptomatic infections. The methods shown here can provide sequencing results within one week of sewage samples arriving in the laboratory. According to the GISAID sequence database [10], only 33 B.1.1.7 sequences had been identified on 16^th^ November 2020 in England, one week after the collection date of our first positive sewage sample. Even in the context of the phenomenal SARS-CoV-2 sequencing program established in England, clinical surveillance requires some time to build up enough data representative of current transmission patterns that is comparable to that provided by ES. In addition, as most clinical testing is based on samples from symptomatic infections, a delay of several days is expected between initial infection and viral sequence data availability due to the time required for symptoms to develop, samples to be collected and PCR/sequencing analyses to be completed. Clinical surveillance is highly dependable on rapid and widespread PCR testing as well as an extensive whole-genome sequencing program. This is currently only happening in a handful of countries so, in most locations, ES could be a major asset for monitoring SARS-CoV-2 virus circulation and identifying VOCs. On the other hand, identification of locally circulating VOCs could be delayed using ES at sites that cover very large populations and therefore, community-based ES targeting smaller populations would increase the sensitivity for detection of such VOCs, e.g. current efforts to track down transmission of B.1.1.7 viruses harbouring mutation E484K in England could be helped by community-based ES in areas where isolation of such viruses are known to have occurred.

## Data Availability

All relevant data are shown in the figures and tables included in the manuscript.

## Acknowledgements

We thank Mihaela Cirdei, Ola Miloszewska, Julian Hand and Maria Zambon from Public Health England for arranging collection and transport of raw sewage samples. This paper is based on independent research commissioned and funded by the NIHR Policy Research Programme (NIBSC Regulatory Science Research Unit). The views expressed in the publication are those of the author(s) and not necessarily those of the NHS, the NIHR, the Department of Health, ‘arms’ length bodies or other government departments.

---

**Table S1.**
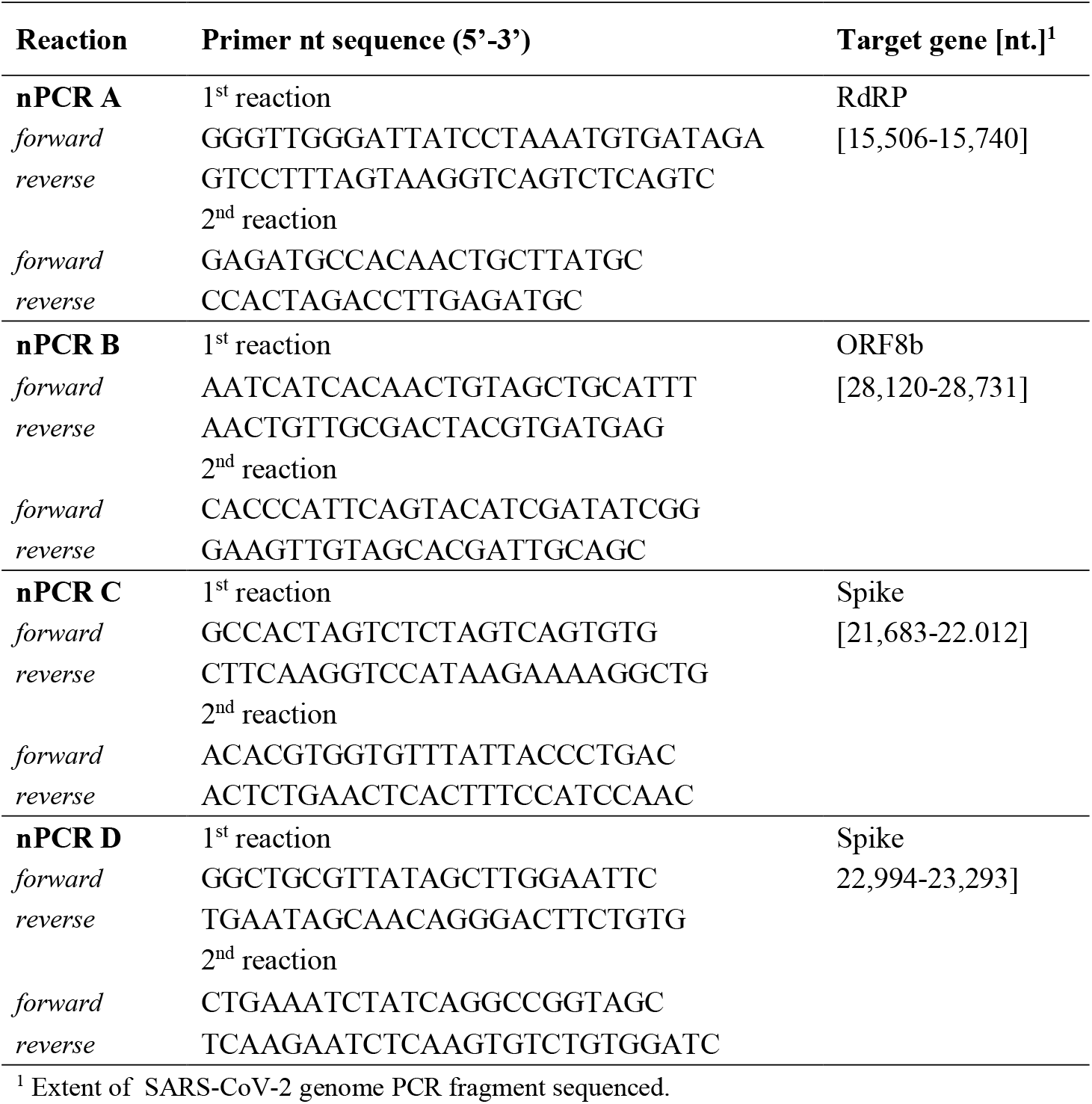
Primers used for nested RT-PCR (nPCR) reactions.

**Figure S1.**
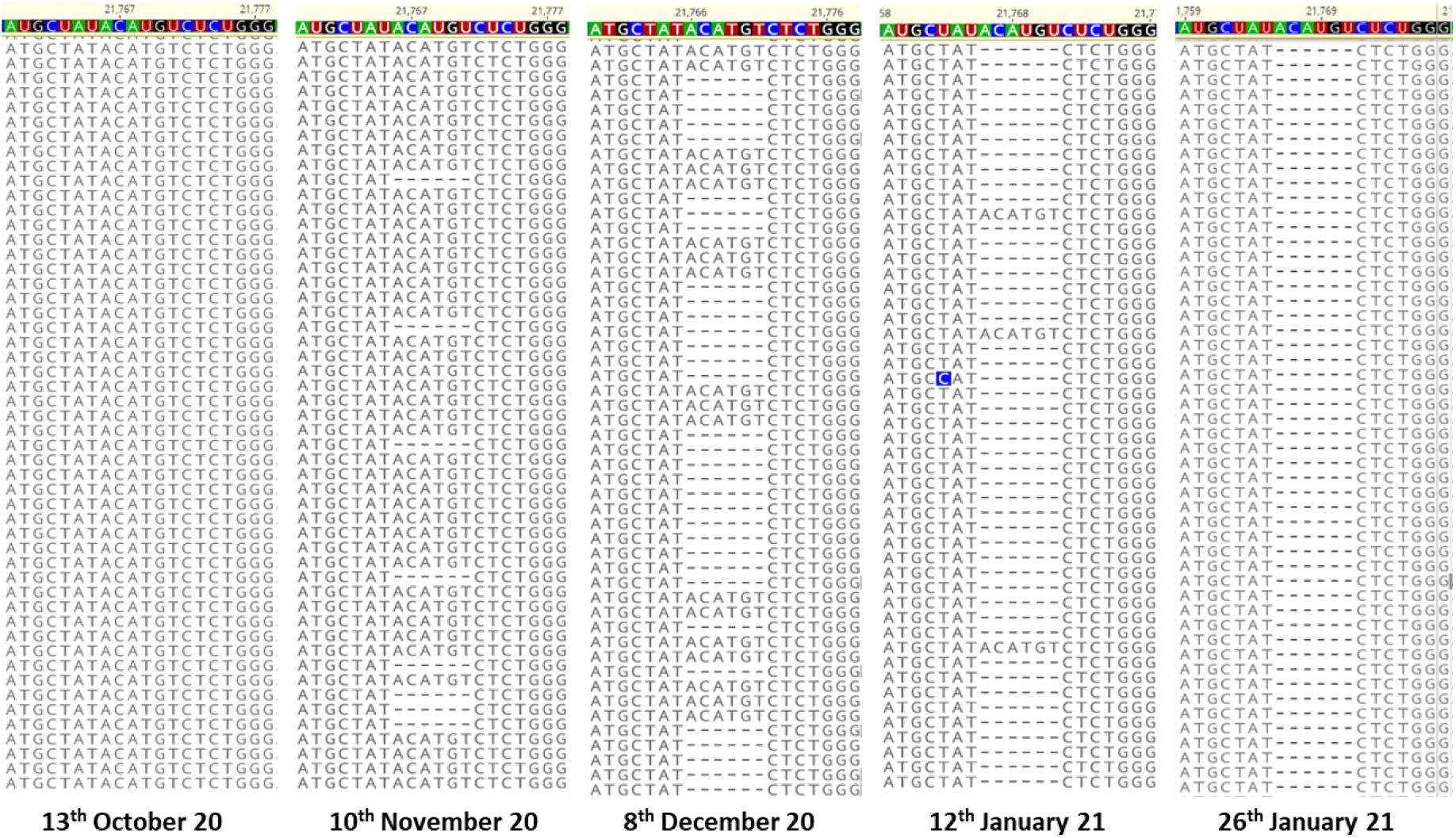
Partial view of NGS contigs showing different proportions of nucleotide 21765–21770 deletion (deletion of amino acids HV69-70) in amplicons from sewage concentraes from different dates. Filtered NGS reads were aligned to SARS-CoV-2 reference sequence (in colour).

## References

1. Chand MH, Susan; Dabrera, Gavin; Achison, Christina; Barclay, Wendy; Ferguson, Neil; Volz, Erik; Loman, Nick; Rambaut, Andrew; Barrett Jeff (21 December 2020). Investigation of novel SARS-COV-2 variant: Variant of Concern 202012/01 (PDF) Investigation of novel SARS-COV-2 Variant of Concern 202012/01. Available at: https://assets.publishing.service.gov.uk/government/uploads/system/uploads/attachment_data/file/959438/Technical_Briefing_VOC_SH_NJL2_SH2.pdf. Accessed 09-02-21 2021.

2. Volz E, Mishra S, Chand M, et al. Transmission of SARS-CoV-2 Lineage B.1.1.7 in England: Insights from linking epidemiological and genetic data. medRxiv 2021.

3. Rambaut A, Loman N, Pybus O, et al. Preliminary genomic characterisation of an emergent SARS-CoV-2 lineage in the UK defined by a novel set of spike mutations. Available at: https://virological.org/t/preliminary-genomic-characterisation-of-an-emergent-sars-cov-2-lineage-in-the-uk-defined-by-a-novel-set-of-spike-mutations/563. Accessed 09-02-21 2020.

4. Chan KK, Tan TJC, Narayanan KK, Procko E. An engineered decoy receptor for SARS-CoV-2 broadly binds protein S sequence variants. bioRxiv 2020.

5. Starr TN, Greaney AJ, Hilton SK, et al. Deep Mutational Scanning of SARS-CoV-2 Receptor Binding Domain Reveals Constraints on Folding and ACE2 Binding. Cell 2020; 182:1295–310 e20.

6. Gu H, Chen Q, Yang G, et al. Adaptation of SARS-CoV-2 in BALB/c mice for testing vaccine efficacy. Science 2020; 369:1603–7.

7. Hoffmann M, Kleine-Weber H, Pohlmann S. A Multibasic Cleavage Site in the Spike Protein of SARS-CoV-2 Is Essential for Infection of Human Lung Cells. Mol Cell 2020; 78:779–84 e5.

8. Johnson BA, Xie X, Kalveram B, et al. Furin Cleavage Site Is Key to SARS-CoV-2 Pathogenesis. bioRxiv 2020.

9. Kemp S, Harvey W, Lytras S, et al. Recurrent emergence and transmission of a SARS-CoV-2 Spike deletion H69/V70. bioRxiv 2021.

10. Elbe S, Buckland-Merrett G. Data, disease and diplomacy: GISAID’s innovative contribution to global health. Glob Chall 2017; 1:33–46.

11. Tegally H, Wilkinson E, Giovanetti M, et al. Emergence and rapid spread of a new severe acute respiratory syndrome-related coronavirus 2 (SARS-CoV-2) lineage with multiple spike mutations in South Africa. medRxiv 2020.

12. Faria NR, Claro IM, Candido D, et al. Genomic characterisation of an emergent SARS-CoV-2 lineage in Manaus: preliminary findings. Available at: https://virological.org/t/genomic-characterisation-of-an-emergent-sars-cov-2-lineage-in-manaus-preliminary-findings/586. Accessed 09-02-2021 2021.

13. Greaney AJ, Loes AN, Crawford KHD, et al. Comprehensive mapping of mutations to the SARS-CoV-2 receptor-binding domain that affect recognition by polyclonal human serum antibodies. bioRxiv 2021.

14. Cheng MH, Krieger JM, Kaynak B, Arditi M, Bahar I. Impact of South African 501.V2 Variant on SARS-CoV-2 Spike Infectivity and Neutralization: A Structure-based Computational Assessment. bioRxiv 2021.

15. Public Health England. Investigation of novel SARS-CoV-2 Variant of Concern 202012/01. Technical briefing 5. Available at: https://assets.publishing.service.gov.uk/government/uploads/system/uploads/attachment_data/file/959426/Variant_of_Concern_VOC_202012_01_Technical_Briefing_5.pdf.

16. Wu K, Werner AP, Moliva JI, et al. mRNA-1273 vaccine induces neutralizing antibodies against spike mutants from global SARS-CoV-2 variants. bioRxiv 2021.

17. Wang Z, Schmidt F, Weisblum Y, et al. mRNA vaccine-elicited antibodies to SARS-CoV-2 and circulating variants. bioRxiv 2021.

18. Wang P, Liu L, Iketani S, et al. Increased Resistance of SARS-CoV-2 Variants B.1.351 and B.1.1.7 to Antibody Neutralization. bioRxiv 2021.

19. Xie X, Liu Y, Liu J, et al. Neutralization of SARS-CoV-2 spike 69/70 deletion, E484K, and N501Y variants by BNT162b2 vaccine-elicited sera. bioRxiv 2021.

20. Muik A, Wallisch AK, Sanger B, et al. Neutralization of SARS-CoV-2 lineage B.1.1.7 pseudovirus by BNT162b2 vaccine-elicited human sera. Science 2021.

21. Foladori P, Cutrupi F, Segata N, et al. SARS-CoV-2 from faeces to wastewater treatment: What do we know? A review. Sci Total Environ 2020; 743:140444.

22. Martin J, Klapsa D, Wilton T, et al. Tracking SARS-CoV-2 in Sewage: Evidence of Changes in Virus Variant Predominance during COVID-19 Pandemic. Viruses 2020; 12.

23. Korber B, Fischer WM, Gnanakaran S, et al. Tracking Changes in SARS-CoV-2 Spike: Evidence that D614G Increases Infectivity of the COVID-19 Virus. Cell 2020; 182:812–27 e19.

24. Fontenele RS, Kraberger S, Hadfield J, et al. High-throughput sequencing of SARS-CoV-2 in wastewater provides insights into circulating variants. medRxiv 2021.

25. Nemudryi A, Nemudraia A, Wiegand T, et al. Temporal Detection and Phylogenetic Assessment of SARS-CoV-2 in Municipal Wastewater. Cell Rep Med 2020; 1:100098.

26. Crits-Christoph A, Kantor RS, Olm MR, et al. Genome Sequencing of Sewage Detects Regionally Prevalent SARS-CoV-2 Variants. mBio 2021; 12.

27. Jahn K, Dreifuss D, Topolsky I, et al. Detection of SARS-CoV-2 variants in Switzerland by genomic analysis of wastewater samples. medRxiv 2021.

28. Izquierdo Lara RW, Elsinga G, Heijnen L, et al. Monitoring SARS-CoV-2 circulation and diversity through community wastewater sequencing. medRxiv 2020.

29. Didion JP, Martin M, Collins FS. Atropos: specific, sensitive, and speedy trimming of sequencing reads. PeerJ 2017; 5:e3720.

30. Office for National Statistics. Coronavirus (COVID-19) Infection Survey, UK: 8 January 2021. Available at: https://www.ons.gov.uk/peoplepopulationandcommunity/healthandsocialcare/conditionsanddiseases/bulletins/coronaviruscovid19infectionsurveypilot/8january2021. Accessed 09-02-2021 2021.

31. Xu C, Wang Y, Liu C, et al. Conformational dynamics of SARS-CoV-2 trimeric spike glycoprotein in complex with receptor ACE2 revealed by cryo-EM. Sci Adv 2021; 7.

